# PREDICTORS OF VIRAL REBOUND AMONG ADOLESCENTS AT AN URBAN CLINIC IN KAMPALA USING REPEATED EVENTS SURVIVAL ANALYSIS

**DOI:** 10.1101/2024.07.29.24311163

**Authors:** Anthony Kirabira, Justine Bukenya, John Ssenkusu, Noah K Ssekamatte, Nazarius M Tumwesigye, Noah Kiwanuka

## Abstract

**Background:** A suppressed HIV viral load below 1000 copies/ml is mark of HIV treatment success because it is associated with reduced risk of transmission of HIV. However, following viral suppression, some people experience viral rebound which may occur multiple times. We used repeated events survival analysis to assess the predictors of viral rebound among adolescents (aged 10-19 years) at an urban clinic in Kampala, Uganda.

**Methods:** The study was a retrospective cohort design conducted at Baylor-Uganda, an HIV care facility. The Lognormal model was used to estimate time from viral suppression to viral rebound (in months) and to determine factors associated with time to first viral rebound. The Prentice, Williams, and Peterson (PWP) model was used to determine the factors associated with repetitive viral rebound.

**Results:** Data from 219 participants were included in the study; 160 (73.06%) were female, and 117 (53.42%) were aged 15-19 years. The overall proportion of viral rebound was 31.5% (31.51/100); 23.29% (23.29/100) experienced one rebound whereas 8.22% (8.22/100) had multiple rebounds. The probability of viral rebound did not reach 50%, so the median time from viral suppression to viral rebound could not be estimated. The 25^th^ percentile survival time to first viral rebound was 34.1 months. The incidence rate of first viral rebound was 84.7 (95%CI, 66.9 - 107.2) per 10,000-person months of observation. The predictors of first viral rebound included; duration on ART (adjusted Time Ratio (TR), 1.04; 95%CI, 1.04-1.05; p<0.001), having psychosocial issues (adjusted TR, 0.67; 95%CI, 0.58-0.77; p<0.001), baseline viral load of <1000 (adjusted TR, 0.85; 95%CI, 0.72-0.99; p=0.008) and protease inhibitors (PI) based ART regimens (adjusted TR, 0.67; 95%CI, 0.49-0.92; p=0.012). The predictors of multiple rebounds included duration on ART (adjusted Hazard Ratio (HR), 0.86, 95%CI, 0.84-0.89; p<0.001), having psychosocial issues (adjusted HR, 11.04, 95%CI, 6.09-20.0; p<0.001), WHO clinical stage II (adjusted HR, 2.28, 95%CI, 1.22-4.25; p=0.002), and WHO clinical stage III (adjusted HR, 2.17, 95%CI, 1.14-4.14; p=0.005)

**Conclusion:** In an urban HIV care facility in Kampala, we found an overall proportion of viral rebound among the adolescents of 31.5%. Occurrence of multiple viral rebounds was associated with duration on ART, psychosocial issues, and WHO clinical staging. Therefore, there is need to incorporate screening of adolescents for psychosocial challenges into the routine programming of HIV care and treatment so as identify and appropriately support those affected in time.

## BACKGROUND

Globally, 38.4 million people were living with HIV at the end 2021. An estimated 0.7% of adults aged 15-49 years worldwide are living with HIV, although the burden of the epidemic continues to vary considerably between countries and regions. Sub Saharan Africa remains the most severely affected, with nearly 1 in every 25 people living with HIV and accounting for more than two thirds of the people living with HIV worldwide (1, 2).

Despite the success in initiating adolescents living with HIV on antiretroviral therapy, maintenance of viral suppression remains a challenge. Previous studies have shown that adolescents with HIV experience suboptimal clinical outcomes when compared to adults (3, 4). This cohort is of importance for research and intervention because adolescents are associated with high rates of new HIV diagnoses each year. Adolescents may experience distinct challenges from older adults such as stigma, which could lead to nonadherence to ART which consequently could increase their chance of experiencing viral rebound after an undetected viral load episode (3, 5).

Viral rebound among adolescents is a great public health phenomenon where single or multiple viral rebounds may occur within the same individual. Failure to maintain viral suppression increases the risk of HIV transmission, occurrence of deleterious clinical outcome (morbidity and mortality) and development of ART resistant strains (6), thereby hampering achievement of the UNAIDS 95–95–95 targets by 2030 (7, 8).

Studies have reported varying levels of HIV viral rebound in low-income countries especially in sub-Saharan Africa. A study in Ghana reported viral rebound of 21% among adult patients on ART whereas a viral rebound rate of 41% was reported in Kenya (7, 9). This indicates that a considerable number of HIV patients who achieved viral suppression were unable to remain suppressed and therefore experienced viral rebound episodes.

According to the Uganda Population HIV Impact Assessment 2022, the prevalence of viral rebound was reported at 24.6% (10). Additionally, a proportion of viral rebound of 29% among adults initiating second line ART in resource limited settings was reported (11). These statistics highlight a significant prevalence of viral rebound among patients receiving ART and yet its associated factors are not well documented.

Over the years, several statistical methods have been developed to handle repeated events such as viral rebound with the ability to adjust for non-independence of events and time between events within an individual. These methods include the extensions of the semi parametric Cox model such as Andersen-Gill (AG), Prentice, Williams and Peterson (PWP), Wei, Lin and Weissfeld (WLW) and the frailty models (12-17). However due to their complex structure and computational requirements, they have not been commonly applied despite the advancement in statistical software in recent years that has made these methods more accessible to researchers (12).

This study therefore applied the different repeated event survival approaches in analysing predictors of viral rebound among adolescents.

## METHODOLOGY

### Study setting

The study was conducted at the Paediatric Infectious Diseases Centre (PIDC)-Mulago also known as Baylor-Uganda. The clinic mainly offers HIV care and treatment to children, adolescents and their caretakers with funding from the Centre for Disease Control (CDC). Over 12,000 clients were enrolled into care at the facility with a range of comprehensive services that include; tuberculosis (TB) management, nutritional support, psychosocial support, and many other child/adolescent friendly services. The centre also partners with the National Institute for Health (NIH) and Medical Research Council (MRC) to conduct research with more focus on clinical trials. The patient records are managed electronically using the EMRx system with a secure data base under the supervision of the data manager.

### Study design

A retrospective cohort study design was adopted for this study. A review of the electronic patient records between 2017 and 2022 from the EMRx system was conducted to identify all the participant information relevant to the study. The study considered adolescents who enrolled on ART between 1^st^ January 2017 and 31^st^ December 2021 and these were followed up until 31^st^ December 2022. The study period therefore, ran from 1^st^ January 2017 to 31^st^ December 2022.

The follow up (time zero) for each participant started from the date a first viral suppression following ART initiation was recorded whereas the follow up ended on the date a viral rebound was recorded.

The gap time for this study was the period from the date a viral rebound was recorded to the date a viral suppression was reported. During this time, an individual was not under follow-up and was also not at risk of rebounding until the date a viral suppression is recorded. (ie an individual only becomes at risk of viral rebound when they report viral suppression)

A single individual had a chance of experiencing more than one viral rebound episode in the study period therefore each participant was followed up until they experienced the first rebound and then given a gap time for them to suppress again before they were followed up for subsequent viral rebounds.

### Study Inclusion/Exclusion criteria

The study population was adolescents aged 10-19 years on ART. Only adolescents aged 10-19 years of age, who newly initiated ART between 1^st^ January 2017 and 31^st^ December 2021 were included into the study and followed up until 31^st^ December 2022. The adolescents aged 10-19 years of age who did not have at least one viral load result of less than 1000copies/ml during the study period were excluded from the study.

### Study Variables

The primary outcome was time from viral suppression to a viral rebound episode. Viral suppression was defined as a patient having viral load measurement of less than 1000copies/ml whereas viral rebound was defined as a viral load result of equal to or more than 1000 copies/ml. The survival time to the first event and between events was calculated as the difference in months from the date of first viral load measurement of less than 1000copies/ml following ART initiation or viral rebound to the date of the first viral load measurement of equal to or more than 1000copies/ml. The survival time of the censored subjects was calculated as the difference in months from the date of the first viral load measurement of less than 1000copies/ml following ART initiation or viral rebound to the date of censoring (i.e. date client is declared lost to follow-up, transferred out, completed study without rebound or death). The event was defined as a participant experiencing viral following viral suppression during the follow up period. A participant was censored when; they were lost to follow-up, transferred out, did not experience any rebound during the study period or died during the study period. This study used right censoring where event free participants at enrolment were followed-up until they either experienced the events or were censored during the follow-up time. The independent variables for the study were baseline viral load, baseline age group, sex, baseline body mass index (BMI), baseline CD4 cell count, baseline WHO clinical stage, TB co-infection, baseline ART regimen, pill number, adherence, duration on ART, drug side effects, dosing schedule, and psychological issues.

### Sampling Procedure

The sampling frame of ART naïve adolescents who initiated ART between 1^st^ January 2017 and 31^st^ December 2021 was generated by the data manager using the clinic patient numbers. The final sampling frame consisted of 219 adolescents which was greater than the actual sample size by only 3 participants. Therefore, the whole 219 participants were considered for this analysis.

### Data Abstraction Procedure

The electronic data abstraction tool was designed by the researcher using Epi-info ver.7 to aid in the extraction of the required information from the EMRx system. The codes of the different categorical variables were incorporated into Epi-info. Research assistants were employed to support the data abstraction process. The research assistants were trained accordingly on how to abstract the data using the Epi-info software. Following successful training of the research assistants, actual abstraction of data for the study started immediately thereafter. The researcher reviewed the extracted data daily and any inconsistencies in the data were identified and rectified by the research assistants every morning before making any new entries.

### Data management

The data set was exported in excel format from Epi-info following completion of the data abstraction process. The copy of the data was backed up on the Google drive. The dataset was imported into STATA 14 where it was checked for completeness, accuracy and consistency. Any inconsistencies identified were rectified. The variable labels were attached to the already coded variables. Variables such as baseline viral load, CD4 cell count, BMI, and age were recategorized to suit the proposed analysis.

The data set was in long format so as to accommodate the recurrent events per individual. This was used in analysis of factors associated with multiple rebounds. The second data set was subset from the multiple rebound data set to include only the first follow-up of each participant. This second data set was used in determining the predictors of first viral rebound following ART initiation.

Multivariate Imputation by Chained Equations (MICE) in R was utilized to handle the missingness. Five data sets were imputed using random forests MICE method following 50 iterations of the MICE algorithm. The Rubin’s rule for pooling parameter estimates for regression models using imputed data sets was not applied because it does not give a log likelihood hence post model estimations such as the AIC and BIC for model comparison could not be obtained.

### Data Analysis

The analyses were done using STATA 14 and R software. Descriptive statistics were obtained for the different variables. Summary statistics particularly the mean and standard deviations, median and the inter-quartile rage (IQR) were obtained for continuous variables whereas frequencies and proportions were generated for categorical variables.

The proportion of single and multiple viral rebounds were obtained through tabulation. Cross tabulations were used to obtain the proportion of single and multiple viral rebounds by sex and age group as well as the mean and standard deviation of number of rebounds.

The Kaplan Meier survival (KM) curves were used to visualize the distribution of survival time to first rebound across the categorical variables mainly sex and age. The median survival time to first viral rebound following ART initiation was computed from the non-parametric KM curve. The log rank test was used to test for the difference in the survival functions of the groups of the different categorical variables. The incidence rate of viral rebound was obtained as well. The assumption of proportionality of hazards was assessed using the proportional hazard plots (-log [-log (St) vs log(t)). Model performance was compared using the AIC and BIC. The model with the lowest AIC and BIC value was considered the best performing model for the data.

The log-normal was the best fitting AFT survival model which was used to determine the predictors of time to first rebound following ART initiation whereas the PWP was best fitting repeated event model that was used to determine the predictors of multiple rebounds. Bivariable analysis was conducted and the variables with a p-value<0.25 were considered for the multivariable regression modelling. The multivariable models were developed using the forward stepwise approach. One variable at a time was added into the model and a variable that had p>0.05 was dropped from the model until parsimony was achieved. Variables with P>0.05 were later re-added into the model to check if they improved model parsimony. Only statistically significant variables (p-value<0.05) were considered the true predictors of viral rebound. The parameter estimates were presented as hazard ratios or time ratios.

### Model diagnostics

The models were assessed for adequacy of fit to the data using the Cox Snell residuals, and the deviance residuals methods and their respective plots were analysed. A plot of Cox Snell residuals that yielded a line that had approximately unit slope or zero intercept suggested that the model may be a good fit of the data whereas for the deviance residuals, any unusual patterns suggested that the model had not adequately fitted the data.

### Ethical considerations

Ethical approval was obtained from the Makerere School of Public Health Institutional Review Board and administrative clearance was obtained from the Research Directorate at Baylor-Uganda. Research assistants were thoroughly trained on the data management protocol to avoid any breach of confidentiality. No personal identifiers were included in the data. Data was not accessible to any third parties besides the study staff. Each study staff had their private login credentials.

## RESULTS

### Baseline Characteristics of the Participants

Table 1 shows a detailed summary of the participants baseline characteristics. The study had a total of 219 participants that were included in the analysis. Of all the participants, 160(73.06%) were female whereas 59(26.94%) were males. Majority 117(53.42%) of the respondents were aged 15-19 years of age. Majority 150(68.49%) did not experience any viral rebound during follow-up, 51(23.29%) experienced one rebound whereas 18(8.22%) experienced more than one viral rebound during follow-up.

**Table 1:** showing baseline characteristics of participants.

| **Variable** | **Frequency** | **Statistic** |
| --- | --- | --- |
| **Duration on ART (months), median (IQR)** | 219 | 45.8 (26.6-66.5) |
| **Number of rebounds, mean (SD; range)** | 219 | 0.42 (0.71; 0-3) |
| **Pill burden (number of pills taken daily), median (IQR)** | 219 | 3 (2-4) |
| **Event status** |  |  |
| Censored | 150 | 68.49% |
| Rebounded | 69 | 31.51% |
| **Rebound status** |  |  |
| No rebound | 150 | 68.49% |
| One rebound | 51 | 23.29% |
| Multiple rebounds | 18 | 8.22% |
| **Age** |  |  |
| 10-14 | 102 | 46.58% |
| 15-19 | 117 | 53.42% |
| **Sex** |  |  |
| Male | 59 | 26.94% |
| Female | 160 | 73.06% |
| **Baseline Viral load** |  |  |
| <1000 | 175 | 79.91% |
| ≥1000 | 40 | 18.26% |
| Missing | 4 | 1.83% |
| **BMI** |  |  |
| Under weight | 88 | 40.18% |
| Normal weight | 101 | 46.12% |
| Over weight | 23 | 10.50% |
| Obese | 4 | 1.83% |
| Missing | 3 | 1.37% |
| **CD4 counts** |  |  |
| ≥200 | 145 | 66.21% |
| <200 | 45 | 20.55% |
| Missing | 29 | 13.24% |
| **WHO clinical stage** |  |  |
| Stage I | 103 | 47.03% |
| Stage II | 61 | 27.85% |
| Stage III | 34 | 15.53% |
| Stage IV | 21 | 9.59% |
| **TB co-infection** |  |  |
| No | 191 | 87.21% |
| Yes | 28 | 12.79% |
| **ART Regimen** |  |  |
| DTG based | 61 | 27.85% |
| PI based | 11 | 5.02% |
| EFV based | 147 | 67.12% |
| **Side effects** |  |  |
| No | 169 | 77.17% |
| Yes | 50 | 22.83% |
| **Dosing schedule** |  |  |
| Once daily | 198 | 90.41% |
| Twice daily | 21 | 9.59% |
| **Psycho-social issues** |  |  |
| No | 152 | 69.41% |
| Yes | 67 | 30.59% |
| **Adherence of the patient** |  |  |
| Good | 147 | 67.12% |
| Poor | 72 | 32.88% |

**NOTE:** SD, standard deviation; IQR, Interquartile range

### Proportion of viral rebound

As indicated in table 1, the overall proportion of viral rebound among the participants was 31.51% (69/219). The proportion of participants who experienced a single viral rebound during follow-up was 23.29% (51/219) whereas the proportion of participants that experienced multiple viral rebounds during follow-up was 8.22% (18/219). The mean number of viral rebounds during follow-up was 0.42 (SD 0.71; range 0-3)

The proportion of participants that experienced single viral rebounds during follow up was slightly higher among males at 27.12% (16/59) compared to females at 21.88% (35/160). Similarly, the proportion of participants that experienced multiple viral rebounds during follow up was higher among males at 11.86% (7/59) compared to females at 6.88% (11/160). When compared across age groups, the proportion of participants with a single viral rebound was higher among those aged 10-14 years at 27.45% (28/102) compared to 19.66% (23/117) among those aged 15-19 years. The proportion of participants with multiple viral rebounds among those aged 10-14 years was 16.67% (17/102) compared to 0.85% (1/117) among those aged 15-19 years

### Median survival time to HIV viral rebound and the incidence rate of HIV viral rebound

Over the follow-up period of the study, the probability of viral rebound did not reach 50%, so the median time from viral suppression to viral rebound could not be estimated. The overall 25^th^ percentile survival time to first viral rebound was 34.07 months. The 25^th^ percentile survival time to first viral rebound among males was 34.07 months compared to 39.23 months among females. However, a median time to first viral rebound of 63.03 months was attained for the males. On the other hand, the 25^th^ percentile survival time to first viral rebound for those aged 10-14 years was 24.53 months which was significantly lower than that of participants aged 15-19 at 67.27months. Unlike those aged 15-19 years, participants aged 10-14 years attained a median survival time to first viral rebound of 62.13 months.

Figure 1 presents the Kaplan-Meier survival plots and the log rank test results during follow up according to sex and age group. The survival time to first viral rebound for both the males and females was not statistically different (p=0.26, by the log rank test). However, those aged 15-19 years were associated with a longer survival time to first viral rebound when compared with those aged 10-14 years (p<0.001, by the log rank test).

**Figure 1:**
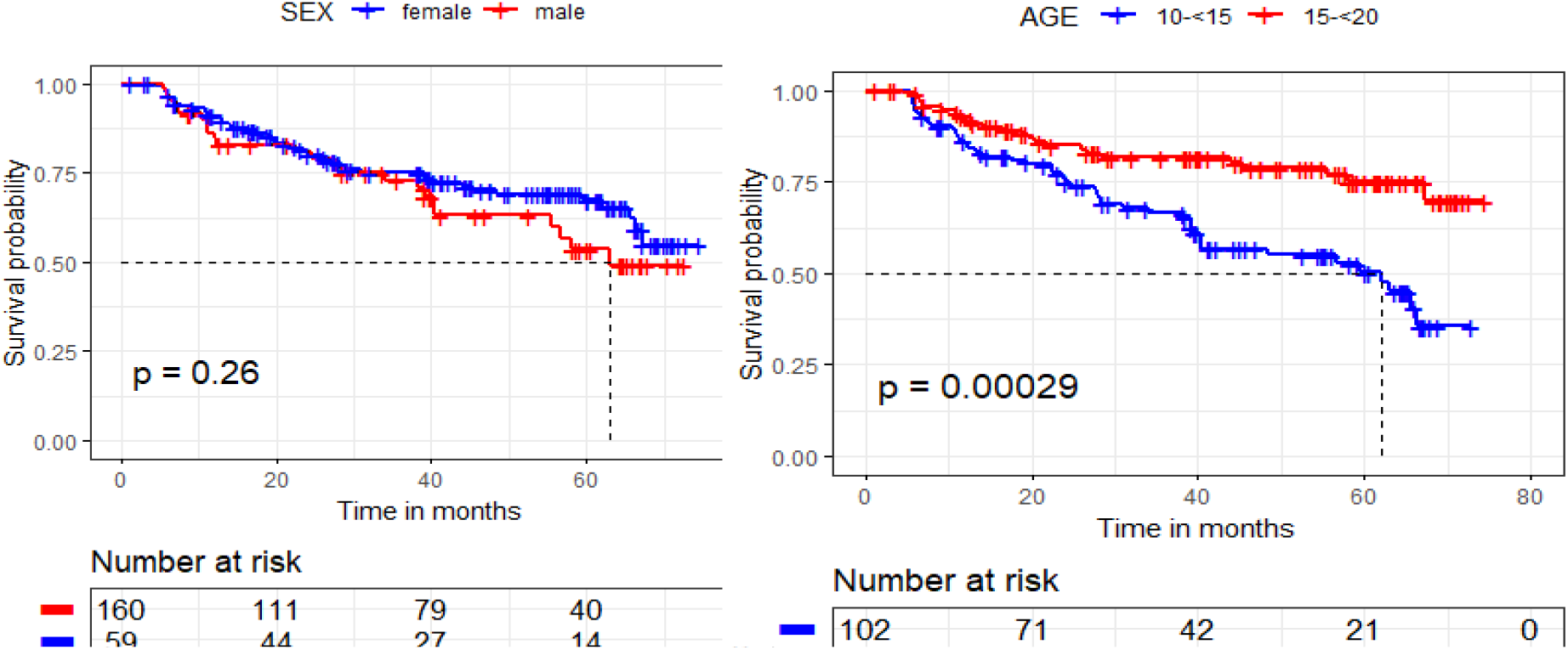
Shows the Kaplan Meier survival plots by age and sex, and their log rank test p-values.

The overall incidence rate of first viral rebound during the 8358.87 person-months of observation was 84.7(95%CI, 66.9-107.2) viral rebounds per 10000-person months of observation. The incidence rate of first viral rebound of 101(95%CI, 69.2-156.7) rebounds per 10000-person months of observation among males was not statistically different from the incidence rate of 76(95%CI, 58.0-103.4) rebounds per 10000-person months of observation among the females. On the other hand, those aged 10-14 years reported a high incidence rate of first viral rebound of 124(95%CI, 92.2-165.5) viral rebounds per 10000-person months of observation. This was more than twice the incidence rate of participants aged 15-19 years at 51(95%CI, 35.7-79.5) viral rebounds per 10000-person months of observation.

### Assessing the PH Assumption

As illustrated in figure 2, the graph of the hazards crossed for two or more sub-groups of different predictor variables mainly sex, ART regimen and CD4 cell count. This confirmed that the PH assumption was violated rendering all semi parametric and parametric PH models not suitable for the analysis. The AFT models (that include; Weibull AFT, Exponential AFT, log logistic, log normal and the generalized gamma model) were hence adopted for the analysis.

**Figure 2:**
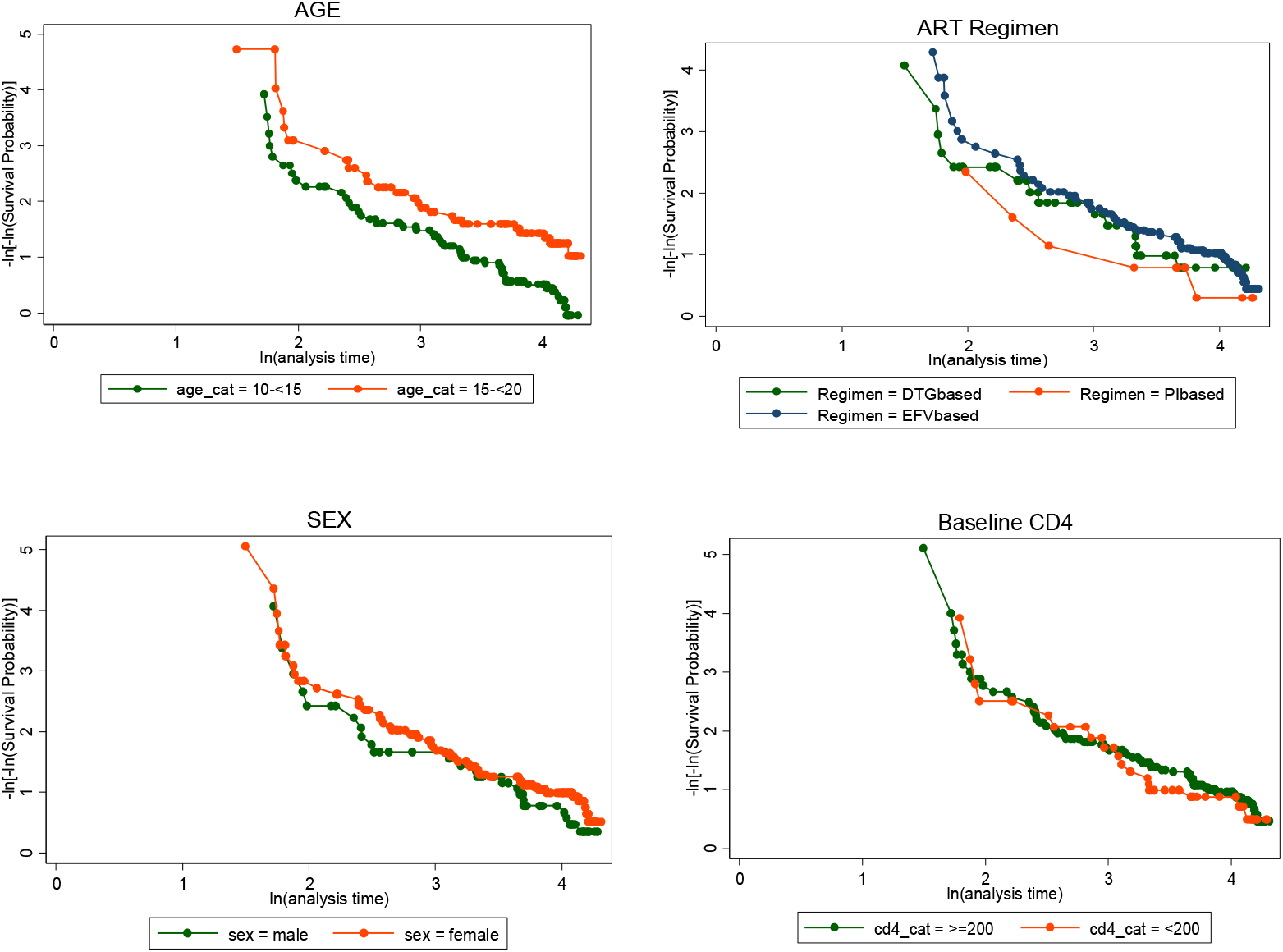
Showing the Log-log plots for checking for the Ph assumption.

### Model comparison

Results presented in Table 2 show that the parametric lognormal model had the lowest value of the AIC and BIC therefore it was considered as the best fit for modelling predictors of time to first viral rebound following ART initiation.

**Table 2:** Showing the AIC and BIC values of the various AFT models.

| Model | AIC | BIC |
| --- | --- | --- |
| Exponential | 242.85 | 290.29 |
| Weibull | 124.34 | 175.18 |
| Lognormal | 122.25 | 173.09 |
| Loglogistic | 124.79 | 175.55 |
| Generalized gamma | 124.21 | 178.41 |

**Table 3:**
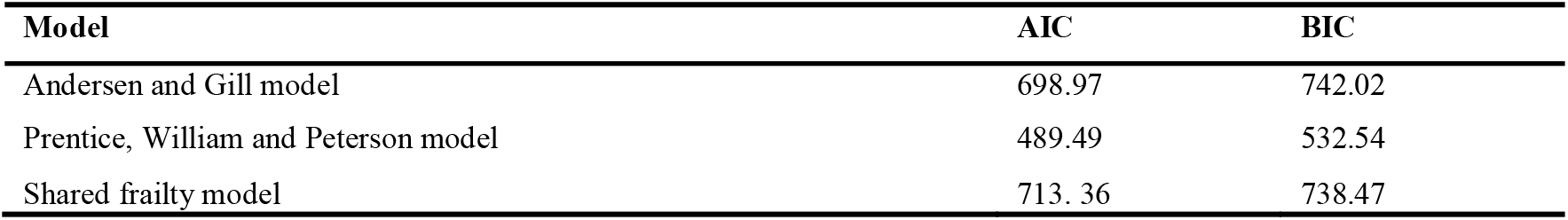
Showing the AIC and BIC of the Various repeated event survival models.

Results presented in table 6 show that the Prentice, William and Peterson (PWP) model had the lowest value of the AIC and BIC. Therefore, the PWP model was considered the best fit in modelling predictors of multiple viral rebounds.

modelling predictors of multiple viral rebounds.

### Predictors of First Viral Rebound following ART Initiation Among Adolescents

Table 4 presents results of the lognormal model that was fitted to analyse the predictors of first viral rebound. Covariates included in the model were age group, sex, duration on ART, pill burden, baseline viral load, body mass index (BMI), CD4 count, WHO clinical stage, TB co-infection, ART regimen, side effects, dosing schedule, and psychosocial issues. The covariates that did not improve model parsimony were dropped however, sex and age group were retained in the final model because viral rebound is known to be associated with age and sex.

**Table 4:** Results of the Lognormal model for the predictors of time to first viral rebound after ART initiation.

| Variable | Unadjusted model estimates |  | Adjusted model estimates |  |
| --- | --- | --- | --- | --- |
|  | TR [95% CI] | P-value | TR [95% CI] | P-value |
| **Age group** |  |  |  |  |
| 15-19 | 1 |  | 1 |  |
| 10-14 | 0.46 [0.29 - 0.75] | 0.002 | 0.93 [0.80 - 1.08] | 0.327 |
| **Sex of Participants** |  |  |  |  |
| Male | 1 |  | 1 |  |
| Female | 1.29 [0.77 - 2.16] | 0.342 | 1.05 [0.90 - 1.22] | 0.524 |
| **Duration on ART** | 1.05 [1.04 - 1.05] | <0.001 | 1.04 [1.04 - 1.05] | <0.001 |
| **Pill burden** | 0.74 [0.64 - 0.86] | <0.001 |  |  |
| **Baseline Viral load** |  |  |  |  |
| ≥1000 | 1 |  | 1 |  |
| <1000 | 0.54 [0.30 - 0.95] | 0.032 | 0.85[0.72 - 0.99] | 0.008 |
| **BMI** |  |  |  |  |
| Normal | 1 |  |  |  |
| Under weight | 0.47 [0.29 - 0.78] | 0.004 |  |  |
| Over weight | 1.56 [0.63 - 3.89] | 0.010 |  |  |
| Obese | 0.40 [0.08 - 2.02] | 0.838 |  |  |
| **CD4_cat** |  |  |  |  |
| ≥200 | 1 |  |  |  |
| <200 | 0.75 [0.44 - 1.27] | 0.284 |  |  |
| **WHO Clinical Stage** |  |  |  |  |
| Stage 1 | 1 |  |  |  |
| Stage 2 | 0.47 [0.27 - 0.84] | 0.010 |  |  |
| Stage 3 | 0.27 [0.14 - 0.52] | <0.001 |  |  |
| Stage 4 | 0.61 [0.26 - 1.42] | 0.253 |  |  |
| **TB Co infection** |  |  |  |  |
| No | 1 |  |  |  |
| Yes | 0.48 [0.25 - 0.91] | 0.025 |  |  |
| **ART Regimen** |  |  |  |  |
| DTG based | 1 |  | 1 |  |
| PI based | 0.80 [0.28 - 2.33] | 0.688 | 0.67 [0.49 - 0.92] | 0.012 |
| EFV based | 1.26 [0.72 - 2.20] | 0.422 | 0.86 [0.73 - 1.02] | 0.083 |
| **Side effects** |  |  |  |  |
| No | 1 |  |  |  |
| Yes | 0.57 [0.33 - 0.97] | 0.038 |  |  |
| **Dosing schedule** |  |  |  |  |
| Once daily | 1 |  |  |  |
| Twice daily | 0.52 [0.25 - 1.08] | 0.078 |  |  |
| **Psycho-social issues** |  |  |  |  |
| No | 1 |  | 1 |  |
| Yes | 0.14 [0.09 - 0.22] | <0.001 | 0.67 [0.58 - 0.77] | <0.001 |

TR= Time Ratios

The variables baseline BMI, WHO clinical stage, TB co infection, pill burden, and side effects were significantly associated with time to first rebound at bivariable analysis however they were not significant at multivariable analysis.

Duration of an adolescent on ART was found to increase the time to first viral rebound after adjusting for age group, sex, baseline viral load, and psychosocial issues (adjusted TR, 1.04; 95%CI, 1.04-1.05; p<0.001). The findings further showed that baseline viral load result of less than 1000copies/ml (adjusted TR, 0.85; 95%CI, 0.72-0.99; p=0.008) and experience of psychosocial issues during follow up (adjusted TR, 0.67; 95%CI, 0.58-0.77; p<0.001) significantly decreased the survival time to first viral rebound following ART initiation after adjusting for duration on ART, sex and age group of the participants. After controlling for other factors, we found that being on protease inhibitors (PI) based ART regimens (adjusted TR, 0.67; 95%CI, 0.49-0.92; p=0.012) resulted into a 33% decrease in the time to first viral rebound compared to those on dolutegravir (DTG) based ART regimens. However, being on efavirenz (EFV) based ART regimens did not have a significant effect on survival time to viral rebound when compared with being on DTG based regimen (adjusted TR, 0.86; 95%CI, 0.73-1.02; p=0.083).

### Model Diagnostics of the Lognormal Model

Figure 3 shows the Cox Snell and deviance residuals of the lognormal model. The Cox Snell residual plot is fairly on a straight line which indicated that the lognormal model fitted the data well. Similarly, the deviance residuals were clustered along the straight line which suggested that the lognormal model still fitted the data well.

**Figure 3:**
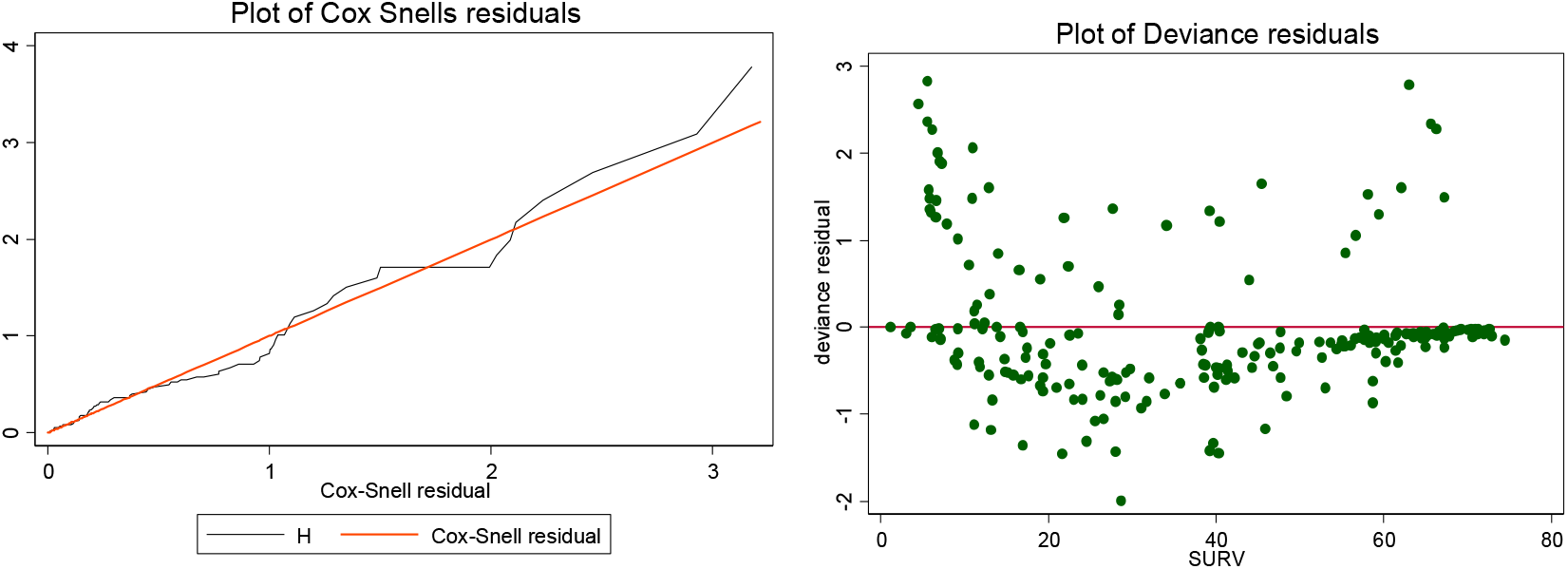
Showing the Cox Snells and deviance residual plots.

### Predictors of Multiple Viral Rebounds Among Adolescents

Table 5 presents hazard ratios (HR) for the effects of covariates on the occurrence of multiple viral rebounds in an individual. At bivariable analysis, the variables age group, pill burden, duration on ART, baseline BMI, WHO clinical stage, TB co infection, side effects and psychosocial issues were significantly associated with occurrence of multiple viral rebounds however, only duration on ART, WHO clinical stage and psychosocial issues remained significant in the final model.

**Table 5:**
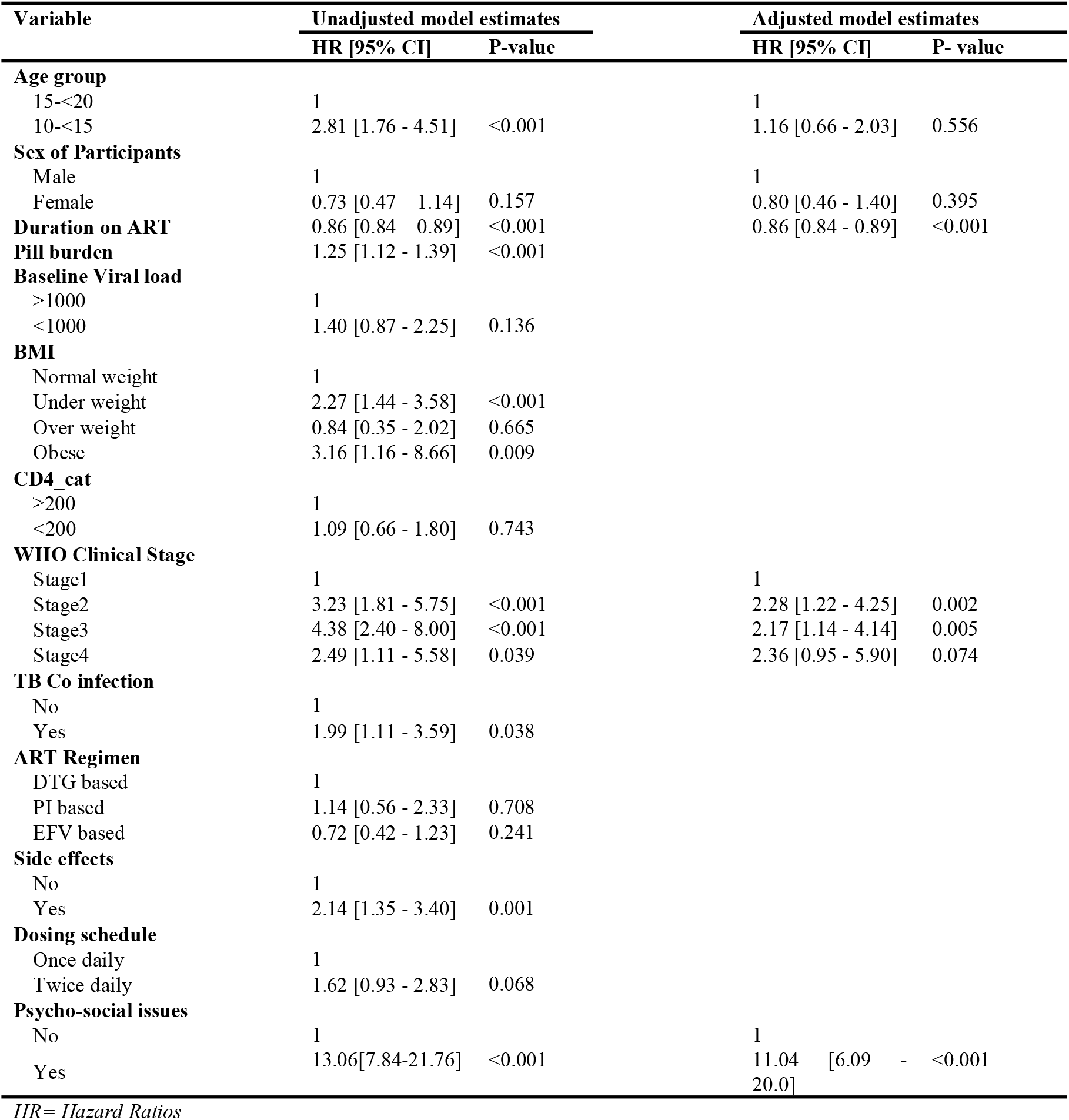
PWP model results of predictors of multiple viral rebounds.

Duration on ART resulted into a decrease in the recurrence of viral rebound while adjusting for other factors (adjusted HR, 0.86, 95%CI, 0.84-0.89; p<0.001). While adjusting for age group, sex, duration on ART, and psycho social issues, we found that adolescents who had WHO clinical stage II (adjusted HR, 2.28, 95%CI, 1.22-4.25; p=0.002) and WHO clinical stage III (adjusted HR, 2.17, 95%CI, 1.14-4.14; p=0.005) were associated with a two-fold increase in the recurrence of viral rebound compared to those who had WHO clinical stage I. There was no significant effect of WHO clinical stage IV (adjusted HR, 2.36, 95%CI, 0.95 - 5.90; p=0.074) on the recurrence of viral rebound when compared with WHO clinical stage I.

Adolescents that experienced psychosocial issues during follow up had an 11-fold increase in the recurrence of viral rebound compared to their counterparts that did not experience any psychosocial issues while adjusting for age group, sex, duration on ART, and WHO clinical stage (adjusted HR, 11.04, 95%CI, 6.09-20.0; p<0.001).

## DISCUSSION

The overall proportion of viral rebound among the adolescents in this study was 31.5% which continues to confirm that a considerable number of patients who achieve viral suppression are unable to maintain it. This proportion is slightly higher than that reported by Palmer et al. (5) and Henri et al., (18) among adolescents in Canada and Central African Republic respectively. Additionally, according to Ng’ambi et al. (19), the proportion of viral rebound decreased with increase in age. This could be attributed to the fact that this is the period young people are transitioning from being dependent to independent. This period is influenced by several social factors that affect adherence such as access to ART, being in schools, denial and stigma.

Additionally, our study found that the proportion of viral rebound was higher among males compared to females. This difference could be explained by the fact that adolescent boys tend to start employment earlier than the girls. This implies that the boys become independent earlier than the girls hence less supervision by the caretakers which increases the risk of non-adherence. This finding is consistent with that of Mpolya (20) who reported a higher viral rebound among males compared to the females.

Although this difference was not statistically significant, it could be of clinical importance.

The median survival time for this study was not attained therefore 25^th^ percentile survival time was reported for this study. The overall 25^th^ percentile survival time to first viral rebound was 34.07 months. This implied that it takes about 34.1 months for 25% of the persons initiated on ART to experience viral rebound. This finding is not consistent with Ng’ambi et al (19) who reported a median survival time to viral rebound of 34.8 months. This discrepancy could be due to the difference in the study periods. The 10-year study period could have enabled the attainment of the median survival time compared to the 6-year study period used in this study.

The overall incidence rate of first viral rebound in this study was 84.7 viral rebounds per 10000-person months of observation. This is higher than the incidence rate of 13.6/10000person months reported by Ng’ambi et al (19) in Malawi. However it’s not consistent with the incidence rate of 183/100person months reported Maina et al., *(9)* in Kenya. Still this could be attributed to the fact that adolescents face a lot of adherence challenges due to the different social factors such as denial, and self-rejection.

The repeated events survival models mainly the AG, PWP, and the shared frailty models were evaluated for better fit in predicting factors associated with viral rebound recurrence using the AIC and BIC. The findings showed that the PWP fitted the data best and was therefore applied in the multivariable analysis of predictors of viral rebound recurrence. This finding is consistent with Thenmozhi et al(21) who reported that the PWP model was found to be better model as compared to other models and also biologically appropriate as subsequent events depend on the previous events.

Every monthly increase in the duration on ART of the adolescents resulted into a 14% decrease in the recurrence of viral rebound. This decrease is attributed to the fact that as the adolescents get older, their acceptance of positive living increases as well which implies better treatment outcomes. This finding is consistent with the findings of Opuku et al. (7) and Maina E et al (9) who reported a similar trend in their studies on viral rebound.

We found that adolescents who had WHO clinical stage II and WHO clinical stage III were associated with a two-fold increase in the recurrence of viral rebound compared to those who had WHO clinical stage I. This finding is consistent with other findings that reported an association between WHO clinical stage II and WHO clinical stage III with multiple viral rebounds Opuku et al. (7).

In this study, adolescents that experienced psychosocial issues during follow up had a 11-fold increase in the recurrence of viral rebound compared to their counterparts who did not experience any psychosocial issues. This could be attributed to the fact that psychosocial issues tend to result into poor adherence hence viral rebound. This finding is consistent with the findings of Gordon (22) who noted psychosocial issues such as belonging to a support group, alive parents and having meals had significant effects on viral rebound.

Its therefore important to evaluate the psychosocial issues of adolescents in line with ART adherence when designing interventions to cub viral rebound.

### Strengths of the study

The study utilized the robust approach of MICE to impute missing data unlike most studies that utilized the complete case analysis approach that tends to decrease the power of the study.

The study also managed to utilize both the parametric AFT models and recurrent events survival models on HIV data which have often has not been applied in the past in HIV research.

### Limitations of the study

The follow-up period of this study was short and therefore the number of rebounds observed in this study was limited by follow-up time of the participants. It could have been possible to experience more rebounds had the follow up time been longer. This would have enabled the attainment of the median time to first viral rebound which was not achieved in this study.

Some of the variables such as distance from facility, being boarding schools, family income, residence, education level, and number of sexual partners present in the conceptual framework were not utilized in the study because they were not captured in the EMRx system at Baylor. These variables had they been included in the models, they could have possibly improved model estimates and changed the conclusion of the study.

## Conclusion

The overall proportion of viral rebound among the adolescents was 31.51%. Occurrence of multiple viral rebounds was associated with duration on ART, psychosocial issues, and WHO clinical staging. Therefore, there is need to incorporate screening of adolescents for psychosocial challenges into the routine programming of HIV care and treatment with focus on adolescents aged 10-14 years.

## Supporting information

This file contains some important tables and figures which could be good for an interested reader

## Data Availability

All data produced in the present study are available upon reasonable request to the authors

