## Supplementary material for "PREDICTORS OF VIRAL REBOUND AMONG ADOLESCENTS AT AN URBAN CLINIC IN KAMPALA USING REPEATED EVENTS SURVIVAL ANALYSIS": This file contains some important tables and figures which could be good for an interested reader

**Table 6: The proportions of viral rebounds by sex and age category**

| **Variable** | **Viral rebound status** | | | |
| --- | --- | --- | --- | --- |
|  | None | Single | Multiple | Total |
| **Sex** |  |  |  |  |
| Male | 36(61.0%) | 16(27.1%) | 7(11.9%) | 59 |
| Female | 114(71.3%) | 35(21.9%) | 11(6.8%) | 160 |
| **Age Group** |  |  |  |  |
| 15 – 19 | 93(79.5%) | 23(19.7%) | 1(0.9%) | 117 |
| 10 -14 | 57(55.9%) | 28(27.5%) | 17(16.7%) | 102 |

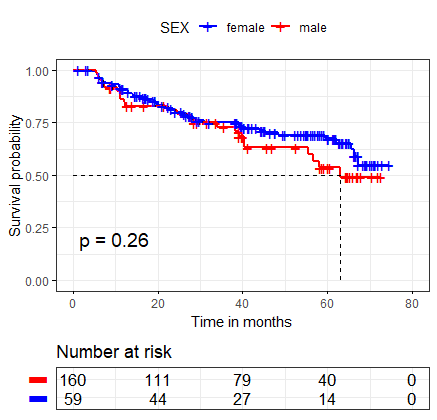

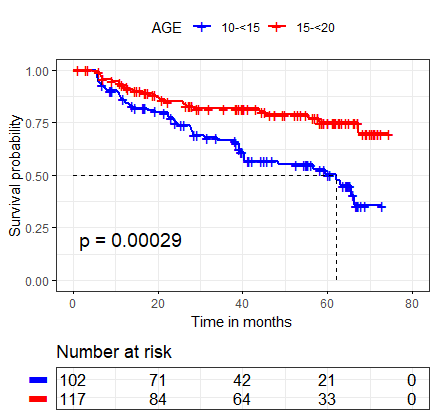

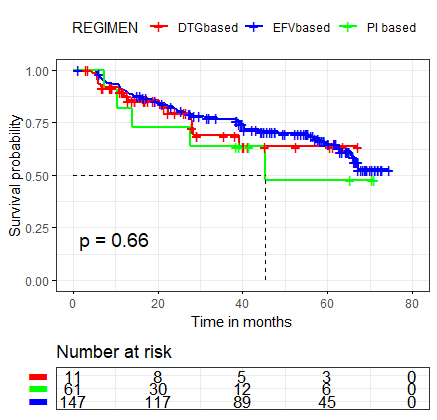

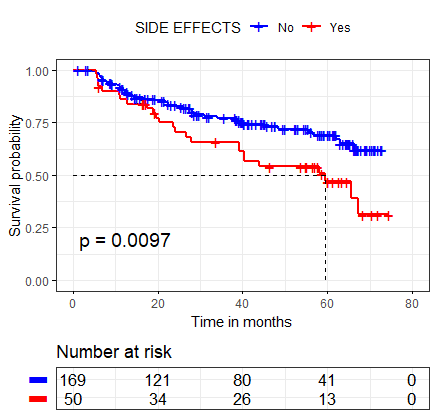

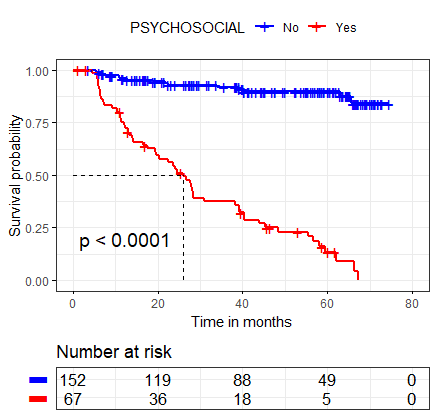

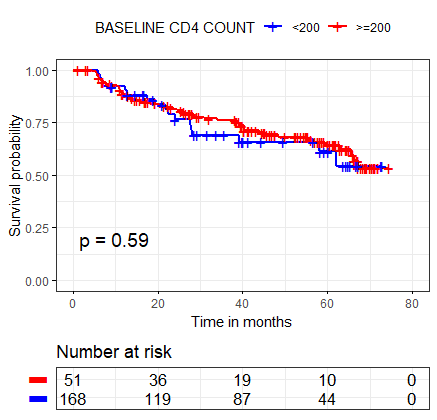

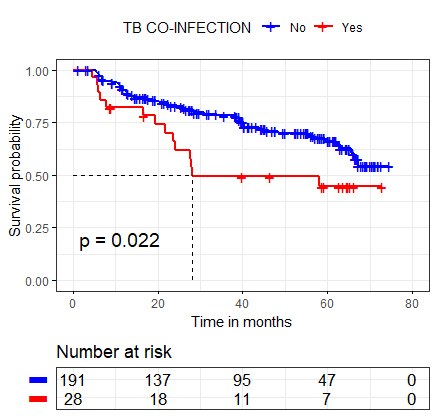

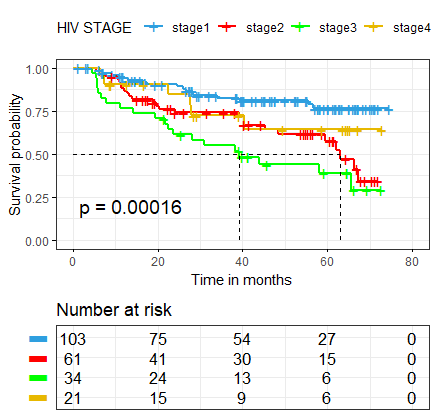

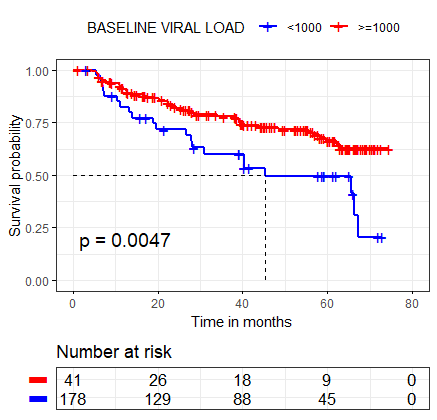

**Figure 4: The Kaplan Meier survival plots and their log rank test p-values**

**Table 7: Summarizing the survival time and incidence rate of first viral rebound**

| **Variable** | **Time at risk (months)** | **Incidence rate per 10000-person months (95% CI)** | **Survival time** | | |
| --- | --- | --- | --- | --- | --- |
|  |  |  | **25th percentile** | **Median** | **75th percentile** |
| **Overall estimates** | 8358.87 | 84.7 (66.9 - 107.2) | 34.07 | NA | NA |
| **Sex** |  |  |  |  |  |
| Male | 2280.17 | 101 (69.2 - 156.7) | 34.07 | 63.03 | NA |
| Female | 6078.7 | 76 (58.0 - 103.4) | 39.23 | NA | NA |
| **Age Group** |  |  |  |  |  |
| 10-14 | 3642.67 | 124 (92.2 - 165.5) | 24.53 | 62.13 | NA |
| 15-19 | 4716.2 | 51 (35.7 - 79.5) | 67.27 | NA | NA |
| **Regimen** |  |  |  |  |  |
| DTG based | 1487.67 | 108 (64.1 - 182.8) | 28 | NA | NA |
| PI based | 430.87 | 116 (48.3 - 278.8) | 14.03 | 45.4 | NA |
| EFV based | 6440.33 | 77.8 (59.0 - 102.7) | 39.23 | NA | NA |
| **Side effects** |  |  |  |  |  |
| No | 6390.33 | 71 (52.8 - 95.4) | 40.4 | NA | NA |
| Yes | 1968.53 | 128 (86.6 - 189.6) | 20.23 | 59.47 | NA |
| **Dosing schedule** |  |  |  |  |  |
| Once daily | 7603.67 | 77 (59.5 - 100.0) | 39.23 | NA | NA |
| Twice daily | 755.2 | 158.9 (90.2 - 279.8) | 22.53 | 40.3 | NA |
| **Psychosocial** |  |  |  |  |  |
| No | 6548.93 | 23.6 (14.2 - 39.2) | NA | NA | NA |
| Yes | 1809.93 | 301 (230.7 - 393.3) | 12.1 | 26 | 48.4 |
| **TB co-infection** |  |  |  |  |  |
| No | 7381.83 | 77 (58.9 - 99.9) | 40.3 | NA | NA |
| Yes | 977.03 | 143 (84.9 - 241.9) | 19.4 | 28 | NA |
| **WHO stage** |  |  |  |  |  |
| Stage1 | 4080.6 | 44 (27.8 - 70.0) | NA | NA | NA |
| Stage2 | 2325.63 | 118 (79.9 - 175.0) | 24.03 | 63.03 | NA |
| Stage3 | 1169.2 | 171 (110.4 - 265.1) | 14.23 | 39.17 | NA |
| Stage4 | 783.43 | 77 (34.4 - 170.5) | 27.97 | NA | NA |
| **BMI** |  |  |  |  |  |
| Under weight | 3209.47 | 136 (100.1 - 184.7) | 22.37 | 59.47 | NA |
| Normal weight | 4027.97 | 55 (36.0 - 82.9) | 45.4 | NA | NA |
| Over weight | 1028.27 | 40 (14.8 - 105.4) | 67.27 | NA | NA |
| Obese | 93.17 | 215 (53.7 - 858.3) | 22.53 | 26.0 | NA |
| **Baseline VL** |  |  |  |  |  |
| Suppressed | 6909.13 | 72 (54.6 - 95.6) | 39.3 | NA | NA |
| Non-suppressed | 1449.73 | 146 (94. 3 - 226.5) | 19 | 45.4 | 67.27 |
| **Baseline CD4** |  |  |  |  |  |
| $\geq$200 | 6534.93 | 79 (60.1 - 104.6) | 39.17 | NA | NA |
| <200 | 1823.93 | 103 (65.8 - 161.8) | 27.67 | NA | NA |

*NA – Not Attained*
